# Prevalence of dementia risk factors in a memory clinic setting: The Oxford Brain Health Clinic

**DOI:** 10.1101/2024.10.01.24314545

**Authors:** J. Blane, G. Gillis, L. Griffanti, R. Mitchell, P.M. Pretorius, S. Forster, S. Shabir, L. Maffei, M.C. O’Donoghue, J. Fossey, V. Raymont, L. Martos, C.E . Mackay

**Author notes:** **<u>Corresponding author:</u>** Jasmine Blane, Oxford Centre for Human Brain Activity, Warneford Hospital, Oxford, OX3 7JX.

## Abstract

With promising disease-modifying therapies (DMTs) emerging and good evidence to support risk reduction in the delay of dementia onset and progression, it is important to understand the profile of patients attending memory assessment services to estimate what proportion of patients might benefit from different types of interventions. The Oxford Brain Health Clinic (OBHC) is a psychiatry-led, clinical-research service that offers memory clinic patients detailed clinical assessments and equal access to research opportunities as part of their secondary care pathway. In this work, we describe the characteristics of OBHC patients in terms of demographics, diagnoses and prevalence of potentially modifiable risk factors compared with a cohort of healthy volunteers and the average memory clinic population. Our results suggest that high research consent rates (91.5%) in the OBHC resulted in a highly representative cohort of the clinical population. Based on Lecanemab trial inclusion criteria, 24.6% of the OBHC population may be suitable for further investigation into DMTs. Furthermore, 67.4% of OBHC patients have at least one potentially modifiable risk factor that may benefit from lifestyle interventions, particularly those focused on depression, sleep and physical activity.

## 1. Introduction

As our population ages, the number of people living with dementia and the prodromal phase, Mild cognitive impairment (MCI), continues to rise, as does the burden on health and social services. It is predicted that by 2050, there will be 1.6 million people living with dementia in the UK, and the economic cost will increase to £47bn^1^. Promising disease-modifying therapies (DMTs), which address the underlying pathology of the diseases that cause dementia, offer hope for change. However, evidence suggests that these treatments are most effective in early-stage Alzheimer’s disease (AD) and due to restrictive exclusion criteria, may be suitable for only a limited number of people with dementia. In a study of community-dwelling adults with a clinical diagnosis of MCI, it was estimated that only 17.4% would be eligible for Lecanemab treatment^2^. Research increasingly supports the involvement of a number of potentially modifiable risk factors in dementia, which, if eliminated, could prevent or delay up to 45% of dementia cases worldwide^3^. The Lancet Commission 2024 listed 14 health and lifestyle factors associated with increased risk of dementia, such as obesity, excessive alcohol consumption and physical inactivity, as well as additional putative risk factors such as sleep duration. The 2023 World Alzheimer Report^4^ focused on dementia risk reduction with the key message ‘never too early, never too late’, which suggests that risk modification could impact a considerable proportion of people already living with MCI and dementia who may not be eligible for DMTs.

In the UK, the diagnosis and management of cognitive impairment in individuals aged over 65 years usually takes place in psychiatry-based memory clinics, but these overburdened National Health Service (NHS) services do not generally have the capacity or means to evaluate risk prevalence, let alone the resources to dedicate to personalised interventions. As a result, much of the research into dementia interventions and risk prevention has taken place in academic settings with bespoke research cohorts that are often distinct from typical memory clinic patient populations. The 2023 national audit of dementia found patients attending Memory Assessment Services (MAS) had a mean age of 79.7 years (range 35-102), with only 6.1% aged under 65 years^5^. By comparison, participants in large dementia cohort studies are typically younger; the Amsterdam Dementia Cohort has an average age of 64 (±10) with 56% aged 65 and under^6^ and participants in the FINGER study, a 2-year multidomain intervention trial targeting a dementia risk factors, were aged 60-77 years^7,8^.

A new type of service is emerging, driven by the prospect of DMTs, the need for risk assessment, communication and personalised risk reduction, and these Brain Health Services (BHS) focus on preventative risk reduction in individuals at high risk of cognitive decline later in life or those with subjective cognitive decline (SCD) or very early, mild cognitive impairment ^9^. Thus, the anticipated population within BHS may be different from current memory clinics, where only 17% of patients receive a diagnosis of MCI and 12.5% receive a non-memory disorder diagnosis or no formal diagnosis^5^. However, to understand which patients may benefit from lifestyle interventions or DMTs, we need to better understand the overall profile of patients attending BHS and current memory assessment services.

The Oxford Brain Health Clinic (OBHC) is a psychiatry-led, clinical-research assessment brain health service for Oxford Health NHS Foundation Trust (OHFT) patients as part of their secondary care, memory clinic assessment^10^. It was launched in August 2020 to assess feasibility, scalability and potential benefits the clinic could offer. Patients complete high-quality clinical assessments that are not routinely available on the NHS and are offered access to research participation, including consenting for their clinical data to be used for research purposes. This service aims to address the gap between clinical practice and research advancements, providing a new mechanism to translate research advancements into clinical settings and create a real-world patient cohort.

The aim of this study was to comprehensively describe the characteristics of the OBHC population in terms of demographics, diagnoses, imaging characteristics and the prevalence of potentially modifiable risk factors. To understand whether the OBHC represents a real-world patient population, we compared the OBHC cohort to healthy volunteers and the average memory clinic population based on 2023 MAS audit^5^. We identified the proportion of patients who may benefit from different types of interventions including DMTs and lifestyle interventions that target potentially modifiable risk factors. The Clarity AD trial inclusion criteria^11^ was used to estimate the proportion of patients who may be eligible for further investigation for Lecanemab. Risk inclusion for this study was based on the life-course model of dementia prevention in the Lancet Commission 2024^3^. We also explored the relationships between risk factor prevalence and cognitive function and subsequent diagnosis.

## 2. Methods

### 2.1. Participants

#### 2.1.1. Clinical population

Patients referred to partner OHFT memory clinics by their general practitioners (GPs) were triaged by a specialty doctor for suitability to attend the OBHC (for more detail, see section 2.1.2 Triaging). At their appointment, all patients were provided with the opportunity to participate in research. Patients could consent to their clinical data collected at the OBHC, as well as relevant data from medical notes, being stored in the OBHC research database, complete additional research assessments during their OBHC visit, and/or choose to hear about future research opportunities. Those who consented to either recontact or additional assessments were required to consent to the use of clinical data for research. All research is optional and patients who chose not to take part in research still complete their NHS assessment at the OBHC.

This paper reports data from participants who attended the OBHC between August 2020 and May 2024 and consented to their clinical data, including subsequent diagnoses, being used for research purposes. The data is stored on the BHC Research Database which was reviewed and approved by the South Central– Oxford C research ethics committee (SC/19/0404).

#### 2.1.2. Triaging

Triaging and decisions regarding suitability to attend the OBHC were made by a specialty doctor and were based on clinical judgment of the need for enhanced assessment and compatibility with the OBHC protocol. This involved reviewing the primary care referral records and patient medical history for possible MRI contraindications or recent scan history, as well as speaking with patients over the phone. All patients requiring imaging are referred to the OBHC unless there is reason to believe that they could not tolerate the visit. The OHBC may have been deemed unsuitable for some patients if there were MRI safety concerns, if the patients were too physically frail for the scan, if they had limited mobility, if they were unable to travel to Oxford, if they did not have an appropriate informant to attend the clinic with them, or if it was felt that cognitive impairment was well established and very advanced; patients may also have elected not to attend the OBHC. Initially, patients who were referred to the OBHC also completed a telephone-based MRI safety prescreening with the radiographer to check for any potential safety concerns; however, this prescreening step was subsequently removed to improve efficiency (patients were still safety screened on the day of their scan). Patients who do not attend the OBHC were referred for a CT scan instead and/or attended the memory clinic appointment as standard.

#### 2.1.3. Healthy volunteers

We compared risk prevalence in our clinical population to a healthy volunteer population obtained from the Brain Health Clinic: Healthy volunteers’ study, which was reviewed and approved by the University of Oxford Medical Sciences Inter-Divisional Research Ethics Committee (R75185/RE001). Data from 81 healthy volunteers aged 65 years and over were included. Participants were recruited from existing research databases and advertisements, with 60.5% from the Join Dementia Research register (National Institute for Health and Care Research), and the remaining participants were recruited from our own databases and from Patient and Public Involvement and Engagement (PPIE) work in our local community, which targeted members of the public who are less research-engaged than those in existing databases. Exclusion criteria included a history of neurological disorders and MRI contraindications.

### 2.2. Procedures

Patients who attended the OBHC completed a series of clinical assessments, including Addenbrooke’s Cognitive Examination (ACE-III)^12^ and 3T MRI brain scans. BMI was calculated from the measured height and weight. Self-report questionnaires were used to collect data regarding educational background, alcohol consumption (Single Alcohol Use Screening Questionnaire^13^), depressive symptoms (Patient Health Questionnaire-9, PHQ9^14^), physical activity (Short Active Lives Survey, SALS^15^), quality of sleep (Pittsburgh Sleep Quality Index, PSQI^14^), long-term health conditions (Long-Term Conditions Questionnaire—short form21, LTCQ-8^16^) and health status (EQ-5D-5L^17^). The questionnaires were posted to the patient ahead of the OBHC appointment to be completed by the patient at home prior to the appointment. Healthy volunteers completed these questionnaires on paper or online at home after their visit.

Additional research assessments the patient could choose to consent to included additional MRI sequences, a saliva sample and for the accompanying relative to complete informant-based research questionnaires. Saliva samples were collected using Oragene OG-600 saliva collection kits (DNA Genotek, Ontario, Canada), and DNA was extracted using Prep-IT-L2P reagent (DNA Genotek, Ontario, Canada) according to the manufacturer’s instructions. Apolipoprotein ε (ApoE) genotyping was performed by LGC Genomics (Hoddesdon, UK). Subsequent patient diagnoses (International Classification of Diseases (ICD-10) codes), and relevant information such as the Bristol Activity of Daily Living Scale^18^, were manually extracted from secondary care electronic healthcare records.

For more details on the assessments performed and the research consent process, please refer to O’Donoghue, et al. ^10^ and Griffanti, et al. ^19^ for the MRI protocol.

#### 2.2.1. Risk Selection

Table 1 shows the included dementia risk factors. As the study aims to determine the proportion of patients who might benefit from lifestyle interventions targeting potentially modifiable risk factors, risk inclusion was based on the potentially modifiable risks highlighted in the Lancet Commission^3^ and the availability of risk data for patients and healthy volunteers. This includes education, alcohol consumption, obesity, depression, and physical activity. As evidence suggests that sleep duration has a U-shaped association with dementia risk^20^, sleep duration was included. Although not modifiable, ApoE genotyping and family history of dementia were included as descriptive characteristics of the patient cohort; ApoE data is not available for healthy volunteers.

**Table 1:** Dementia risk factors included in the analysis.

| Risk | Measure | Details |
| --- | --- | --- |
| Education | Self-report questionnaire | Highest level of education achieved. None/non-secondary level schooling considered high risk. |
| Alcohol Consumption | Self-report questionnaire (MSASQ) | How many times an individual had six or more drinks on a single occasion. High risk is considered >21 drinks per week, which for this questionnaire is equivalent to six or more drinks daily or almost daily. |
| Obesity | BMI | Calculated from measured height and weight. Those considered at high risk have a BMI over 30 (obese). |
| Depression | Self-report questionnaire (PHQ9) | Based on PHQ9 total score. Score of 0-4 Minimal depression, 5-9 Mild depression, 10-14 Moderate depression, 15-19 Moderately severe depression, 20-27 Severe depression. Those considered high risk are those who have moderate-severe depressive symptoms (total score of >9). |
| Physical Inactivity | Self-report questionnaire (SALS) | Activity is split into three categories: walking, cycling and sport/fitness. Participant's total score is determined by calculating the sum of individual activity scores, which are calculated by number of days the activity was completed multiplied by the minutes spent on each activity where it was sufficient to raise breathing rate. Activities which were not sufficient to increase breathing rate are excluded from the calculation. Based on total score, participants are classified as 'active' if they achieve the recommended levels of at least 150 minutes of moderate intensity physical activity per week, 'fairly active' for those between 30-149 minutes per week and 'inactive' for those who are active for less than 30 minutes of activity per week. The 'inactive' group are considered high risk. |
| Sleep | Self-report questionnaire (PSQI) | Self-reported hours of actual sleep at night. Less than 5 hours is considered 'inappropriate', more than 5 hours and less than 7 is 'uncertain', more than 7 and less than 9 hours is 'appropriate', and 10 or more is considered 'inappropriate'. High risk is classified as inappropriate sleep duration (<than 5 or >10 hours of sleep). |
| Family history of dementia | Self-report | Patient data regarding family history of dementia has been extrapolated from medical records which was self-reported at subsequent memory clinic appointments. Data from healthy volunteers was provided via a self-report questionnaire to capture first and second-degree family history of dementia. Those considered at high risk have reported any family history of dementia. |
| ApoE | Extracted from DNA from saliva samples | The three main APOE alleles ($\epsilon 2$, $\epsilon 3$, $\epsilon 4$) were defined based on the combination of variants at two single nucleotide polymorphisms (SNPs) of the APOE gene: *rs429358* and *rs7412*. Depending on the combination of alleles at *rs429358* and *rs7412* variants, six common APOE genotypes can be derived: $\epsilon 2/\epsilon 2$, $\epsilon 2/\epsilon 3$, $\epsilon 2/\epsilon 4$, $\epsilon 3/\epsilon 3$, $\epsilon 3/\epsilon 4$, $\epsilon 4/\epsilon 4$. Presence of an $\epsilon 4$ allele is considered high risk. |

### 2.3. Statistical analysis

Characteristics of the OBHC cohort have been expressed as percentages and frequency counts for categorical variables and means, standard deviations and ranges for continuous variables. The prevalence of a dementia risk factor was defined as the frequency (percentage) of high-risk factors (as per Table 1). Independent samples t-tests were used to explore differences in demographic variables and risk factor prevalence between OBHC patients and healthy volunteers. Where parametric assumptions of normality and homogeneity of variance cannot be met for t-tests, the Wilcoxon rank (Mann-Whitney) nonparametric test was employed. Spearman’s correlation coefficient was used to explore the relationship between age and total ACE-III score in patients and healthy volunteers. Categorical risk variables were dichotomised into the presence of a high-risk factor and the absence of a high-risk factor (as per Table 1), and Pearson’s chi-square tests were used to explore differences in risk prevalence between populations. Where possible, prevalence rates have been compared to average rates reported in the 2023 MAS audit^5^.

To evaluate associations between risk prevalence and diagnosis, primary diagnoses were categorised as dementia-related diagnoses (DRD; ICD-10 codes: F00, F01, F02, F03), mild cognitive impairment (MCI; F06.7), and non-memory disorder diagnoses/no formal diagnoses (no DRD; F10, F31, F32, F41, and patients who received no formal diagnosis) and compared to a cohort of healthy volunteers. ANOVA was used to compare age across diagnostic groups. Parametric assumptions (normality and/or homogeneity of variance) were violated for all continuous risk scores, including PHQ9 total score, SALS total score, sleep duration and BMI as well as cognition (ACE-III total score); consequently, the Kruskal-Wallis test was employed to explore associations between risk prevalence, cognition and diagnostic group.

To determine the proportion of patients that may be eligible for further investigation into DMTs, DMT eligibility was based on inclusion criteria from the CLARITY AD trial of Lecanemab^11^. This includes a diagnosis of MCI (ICD-10: F06.7) or AD (ICD-10: F00.1, F00.0, F00.9, F00), aged 50-90 years and a BMI greater than 17 and less than 35. The trial inclusion criteria also included a Mini-Mental State Examination (MMSE) score >22 and <30. Although the MMSE was not included in the OBHC, using a conversion table^21^, ACE-III scores >63 and <100 were taken as a proxy measure of cognition in place of the MMSE. The CLARITY AD trial^11^ exclusion criteria included a Geriatric Depression Scale (GDS) score of more than 8, indicating moderate depression; while the OBHC did not include the GDS, depressive symptoms were assessed with the PHQ9, for which a score greater than 9 indicates moderate depression. Imaging-based exclusion criteria were evaluated from radiology reports, including more than 4 microhaemorrhages, previous macrohaemorrhage, evidence of superficial siderosis or vasogenic oedema, multiple lacunar infarcts or stroke involving a major vascular territory, severe small vessel (calculated in the OBHC by a Fazekas periventricular white matter hyperintensities or deep white matter hyperintensities score of 3 or more), or other major intracranial pathology. Information on the remaining trial inclusion/exclusion criteria, such as concomitant medication use or exclusionary medical conditions, was not available in the OBHC at the time of analysis.

Total risk prevalence was calculated by the sum of individual risk factor prevalence; ApoE and family history of dementia risk factors were excluded from total risk prevalence as these factors are not modifiable. The Wilcoxon rank (Mann-Whitney) nonparametric test was used to explore differences in total risk prevalence between OBHC patients and healthy volunteers, and the Kruskal-Wallis test was used to explore associations across diagnostic groups. Total risk was then grouped into 0, 1 and 2+ risk factors, and the Kruskal-Wallis test was used to explore associations with cognition (ACE-III total score).

Analyses were conducted using R 4.1.3., p values were two-sided, and statistical significance was defined as p < .05. The availability of descriptive characteristics and risk factors for both the OBHC and healthy volunteer populations are included throughout. In line with clinical practice, we did not impute missing data.

## 3. Results

### 3.1. Cohort demographics of patients and healthy volunteers

A total of 342 patients attended the OBHC between August 2020 and May 2024. Research uptake was high (see Table 2.); 91.5% (n=313) consented to their clinical data, including subsequent diagnosis, being used for research purposes. 71.4% of patients consented to be contacted about future research compared to 12.8% in the MAS 2023 audit^5^.

**Table 2:** Uptake of research by the OBHC patient population. Number and percentage of patients who consented to join the BHC research database, to complete additional assessments and to be contacted in the future for other research opportunities.

|  | Use of clinical data for research | Additional research assessments |  |  |  | Recontact about future research |
| --- | --- | --- | --- | --- | --- | --- |
|  |  | Any additional assessment | MRI | Saliva | Informant questionnaire |  |
| **Consented, N (%)** | 313 (91.5%) | 294 (86.0%) | 230 (67.3%) | 267 (78.1%) | 284 (83.0%) | 244 (71.4%) |
| *% of total patient attendees, n= 342* |  |  |  |  |  |  |

Table 3. shows the demographic and diagnostic characteristics of 313 patients and 81 healthy volunteers. 54.3% of the OBHC population was male, whereas 44.4% of the healthy volunteers were male; this difference was not significant (X^2^(1) =2.51, p=.11; Fisher exact: p=.13). Of those whose native language was known, 95.4% of patients spoke English as a first language compared to 97.3% of healthy volunteers (X^2^(1) =0.48, p=.49; Fisher exact: p=.17). Of the OBHC patients whose ethnicity was known, 99.0% were white. By comparison, the 2023 MAS audit found that 43% of patients attending memory assessment services were male, 91.6% of patients whose native language was known spoke English as a first language, and 87.9% of those whose ethnicity was known were white^5^.

**Table 3:** Demographics and diagnoses (ICD-10) breakdown for OBHC patients, healthy volunteers, and the average memory clinic population per the Memory Assessment Service (MAS) audit^5^.

| Sample Characteristic | OBHC Patients | Healthy volunteers | MAS 2023<sup>5</sup> |
| --- | --- | --- | --- |
| **Demographics (n)** | 313 | 81 | 5899 |
| Age (years) - mean ± s.d (range) | 77.9 ± 6.3 (65-101) | 76.4 ± 5.9 (65-92) | 79.7 (35-102) |
| Sex (Male/Female) | 170/143 | 36/45 | 2642/3506 |
| Lacked capacity (%) | 15.4% | 0% | n/a |
| Ethnicity (White/non-white/unknown %) | 31.4/0.3/68.6% | n/a | 77.2/10.7/12.2% |
| English as first language (Yes/No/Unknown %) | 93.9/4.5/1.6% | 87.7/2.5/9.9% | 86.2/7.9/6% |
| **Cognitive Scores (n)** | 302 | 81 | n/a |
| ACE-III total score (max 100) - mean ± s.d. (range) | 74.4 ± 17.3 (9-99) | 95.4 ± 4.3 (78-100) | n/a |
| ACE-III memory sub-score (max 26) - mean ± s.d. (range) | 16.0 ± 6.6 (0-26) | 24.7 ± 2.1 (17-26) | n/a |
| **Other (n)** | 237/143/94/313 | 40 | n/a |
| LTCQ-8 - mean ± s.d. (range) | 72.5 ± 15.9 (21.9-100) | 85.7±18.2 (31.3-100) | n/a |
| BADLS - mean ± s.d. (range) | 10.4 ± 11.1 (0-51) | n/a | n/a |
| EQ-5D-5L visual analogue score - mean ± s.d. (range) | 80.3 ± 18.1 (0-100) | n/a | n/a |
| Clinical Frailty Scale - mean ± s.d. (range) | 3.0 ± 1.3 (1-7) | n/a | n/a |
| **Diagnosis ICD-10 (n)** | **286** | **n/a** | **6148** |
| AD, late onset [F00.1] (n) | 28.3% (8) | n/a | 30.1% (1848) |
| AD, early onset [F00.0] (n) | 1.7% (5) | n/a |  |
| AD, unspecified [F00.9/F00] (n) | 2.1% [0.7/0.4%]<br>(6 [2/4]) | n/a |  |
| AD, mixed [F00.2] (n) | 13.3% (38) | n/a | * |
| F01 Vascular dementia<br>[F01/F01.1/F01.2/F01.3/F01.9] (n) | 3.5%<br>[0.3/0.3/0.3/1.4/1.0%]<br>(10 [1/1/1/4/3]) | n/a | 11.9% (729) |
| F02 Dementia in other diseases classified elsewhere [F02/F02.3/F02.8] (n) | 1.7% [0.3/1.0/0.3%]<br>(5 [1/3/1]) | n/a | 3.7% (230) |
| Unspecified Dementia [F03] (n) | 1.7% (5) | n/a | 4.0% (248) |
| Mild Cognitive Impairment [F06.7] (n) | 26.9% (77) | n/a | 17% (1045) |
| F31 Bipolar disorders [F31.3/F31.7] (n) | 0.7% [0.3/0.3%]<br>(2 [1/1]) | n/a | 1% (63) |
| F32 Depressive Episode [F32.0/F32.1] (n) | 1.4% [0.3/1.0%]<br>(4 [1/3]) | n/a |  |
| F41 Anxiety Disorders [F41.1/F41.2] (n) | 1.0% [0.7/0.3%]<br>3 (2/1) | n/a |  |
| No formal diagnosis/subjective cognitive impairment/other (not dementia) (n) | 17.5% (50) | n/a | 11.5% (705) |

When categorised by diagnosis, the largest proportion of OBHC patients received a dementia related diagnosis (52.4%, n=150), followed by a diagnosis of MCI (26.9%, n=77), and the smallest proportion received a non-memory disorder diagnosis or no formal diagnosis (20.6%, n=59). Compared with national rates, the OBHC had a lower percentage of patients with a dementia-related diagnosis and a higher proportion of patients with MCI and non-memory disorder diagnosis/no formal diagnosis (MAS audit: 70.5% dementia, 17% MCI and 12.5% non-dementia^5^). Among the OBHC dementia subgroup, 86.7% had a diagnosis of dementia in Alzheimer’s disease (F00) and 13.3% had a non-AD dementia diagnosis; compared to 42.6% and 57.4% of patients nationally^5^.

Figure 1 shows the age and ACE-III total score distributions and correlations for the OBHC patient and healthy volunteer populations. The age distributions of OBHC patients (77.9 ± 6.3 years, range: 65-101) and healthy volunteers (76.4 ± 5.9 years, range: 65-92) were similar, with a no significant difference in age between the groups (t(132.66)=1.69, p=.09 with small effect size d=0.20). Most patients and healthy volunteers are aged between 70 and 85 years. By comparison, the mean age of patients attending MAS was 79.7 years (range 35-102), with 93.9% aged 65 years and older^5^.

**Figure 1.**
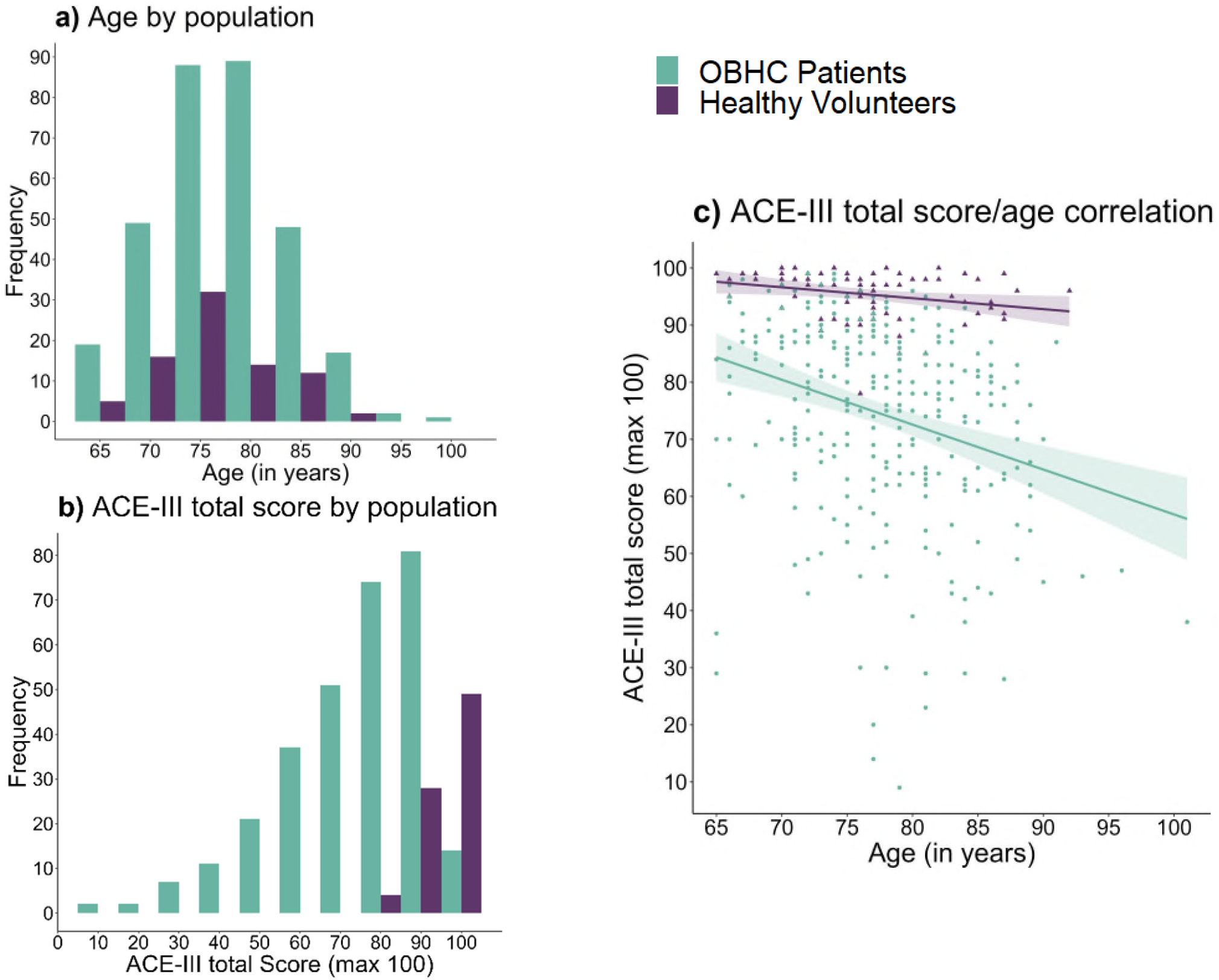
Demographic comparison of OBHC pafients (green) and healthy volunteers (purple) including a) Age distribufion. b) ACE-III total score distribufion. c) Age and ACE-III total score correlafion. ACE-III = Addenbrooke’s Cognifive Examinafion-III.

Compared with healthy volunteers, OBHC patients scored significantly lower on the ACE-III in both the total score (W=1648.5, p<.001, effect size=-0.61) and memory sub-score (W=2326, p<.001, effect size =-0.57). 78.3% of patients (n=235) had a total ACE-III score of 88 or lower, and 58.7% (n=176) had a total ACE-III score of 82 or lower; in comparison, 6.2% (n=4) of healthy volunteers had a total score of 88 or lower, and 1.2% (n=1) had a total ACE-III score of 82 or lower. There was a significant negative correlation between age and the ACE-III total score for both patients (rs=-.32, p<.001) and healthy volunteers (rs=-.33, p =.003); the difference in these correlations was not significant (p=.49).

### 3.2. Imaging characteristics

92.7% of OBHC patients completed the full clinical MRI scan, and 54.6% completed the full research MRI scan; 97.5% of healthy volunteers completed the full clinical and research scans. By comparison, the MAS audit disclosed that brain imaging (MRI or CT) was requested for 47.3% of patients (n=2910), completed for 44.3% (n=2725) and 13.2% (n= 812) had a MRI scan; the most common reason that the requested scans were not performed was patient decline (40.5%), previous scan (15.1%), not required (7.6%) and contraindication (1.1%); the reason was unknown for 35.7% of the patients. Table 4 shows the imaging characteristics of the OBHC patients from currently available radiology reports.

**Table 4:** Summary of completed clinical and research MRI scans (n) and main findings from radiology reports.

|  |  |
| --- | --- |
| **Clinical sequences (n)** | **299** |
| Fully completed | 290 |
| Partially completed | 9 |
| **Research sequences (n)** | **181** |
| Fully completed | 171 |
| Partially completed | 10 |
| **Radiology report (n)** | **275** |
| Generalised atrophy (counts for none/mild/moderate/severe) | 11/170/91/3 |
| L MTA score – median (range) [counts] | 2 (0–4) [7/76/88/84/16] |
| R MTA score – median (range) [counts] | 2 (0–4) [7/79/90/77/18] |
| Average MTA score - median (range) [counts] | 2 (0–4) [7/70/72/68/12] |
| Fazekas PWMH score – median (range) [counts] | 1 (0–3) [11/129/90/39] |
| Fazekas DWMH score – median (range) [counts] | 1 (0–3) [16/155/88/11] |
| Fazekas total score – median (range) [counts] | 3 (0–6) [3/21/106/47/55/27/10] |
| Microhaemorrhages (yes/no) | 43/229 |
| Previous haemorrhages (yes/no) | 12/7 |
| Previous chronic or acute infarcts (yes/no) | 2/173 |
| Restricted diffusion suggesting prion disease (yes/no) | 0/272 |
| Mass (yes/no) | 13/260 |
| Extra-axial collection (yes/no) | 0/271 |
| Hydrocephalus (yes/no) | 0/272 |
| *MTA = Medial Temporal lobe Atrophy; PWMH = periventricular white matter hyperintensities; DWMH = deep white matter hyperintensities.* |  |

### 3.3. DMT eligibility

To explore the proportion of patients who may be eligible for DMTs, we calculated potential eligibility for Lecanemab on the basis of the CLARITY AD trial^11^ inclusion/exclusion criteria as the first treatment for AD licenced for use in Great Britain by the Medicines and Healthcare Products Regulatory Agency (MHRA). This includes diagnosis, age, cognition, BMI, depressive symptoms, and MRI exclusion criteria based on radiology reports (for more detail, see section 2.3. Statistical analysis). 75.4% of the OBHC population did not meet the trial eligibility criteria, with 149 patients excluded based on age and diagnosis only and an additional 81 excluded based on cognition, BMI, depressive symptoms, and imaging characteristics. Consequently, 24.6% of patients in the OBHC population may be eligible for further investigation into the Lecanemab trial criteria, including exploration of brain amyloid pathology.

### 3.4. Risk Prevalence

Compared with healthy volunteers, OBHC patients tended to have a greater prevalence of potentially modifiable dementia risk factors, and the associations between risk prevalence and population were statistically significant for depressive symptoms (PHQ9; X^2^(1) = 5.92, p=.015; Fisher exact: p=.015), physical activity (SALS; X^2^(1) = 16.30, p<.001; Fisher exact: p<.001) and sleep (PSQI; X^2^(1) = 5.61, p=.017; Fisher exact: p =.016), with a greater presence of categorical “high risk” factors in patients than in healthy volunteers.

Figure 2 shows the prevalence of potentially modifiable dementia risk factors in the OBHC patient population. 16.7% (n=49) of OBHC patients reported moderate-severe symptoms of depression on the PHQ9, compared to 5.5% (n=4) of healthy volunteers. 54.2% (n=169) of OBHC patients and 27.8% (n=20) of healthy volunteers reported engaging in less than 30 minutes of moderate-intensity physical activity per week and 15.9% (n=47) of OBHC patients and 8.6% (n=7) of healthy volunteers had a BMI over 30, indicating obesity. 8.3% (n=23) of OBHC patients reported no formal qualifications compared to 4.6% (n=3) of healthy volunteers, 1.0% (n=3) of OBHC patients and 0% of healthy volunteers reported consuming 6 or more drinks on a single occasion daily or almost daily, and 10.5% (n=32) of OBHC patients compared to 1.5% (n=1) of healthy volunteers reported an inappropriate sleep duration of less than 5 or more than 10 hours per night.

**Figure 2.**
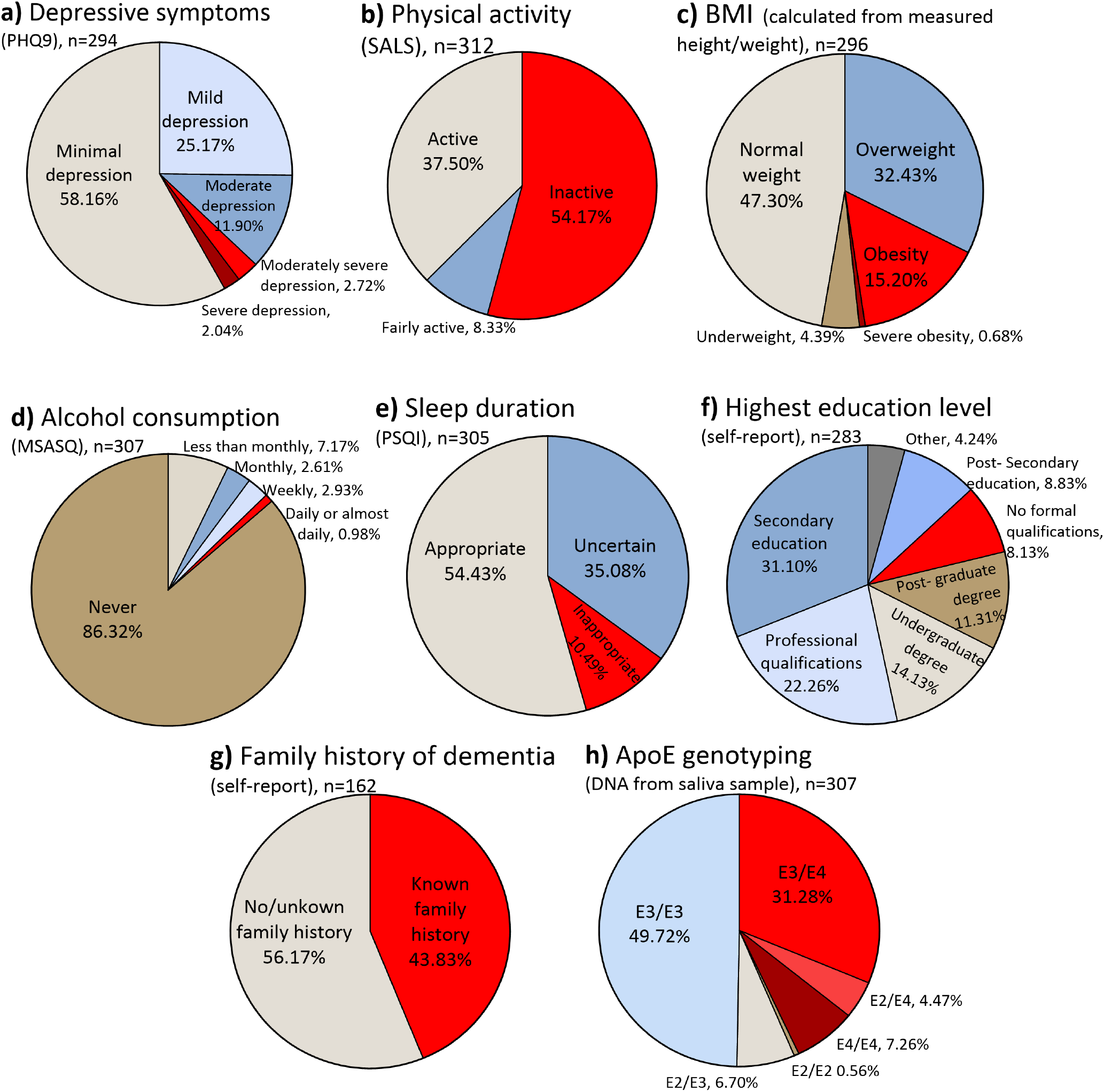
Prevalence of potenfially modifiable demenfia risk factors and non-modifiable risk factors in the OBHC pafient populafion. Modifiable risk factors include depressive symptoms, physical acfivity, BMI, alcohol consumpfion, sleep durafion, highest educafion level. Non-modifiable risk factors include ApoE genotype and family history of demenfia. High risk factors are shown in red/dark red.

Self-reported, known family history of dementia was available for 162 OBHC patients and 72 healthy volunteers; 43.8% of patients and 52.8% of healthy volunteers had a known history of dementia; however, patient and heathy volunteer data was collected using different methods. ApoE genotyping was available for 180 OBHC patients, of whom 42.8% (n=62) had at least 1 ApoE ε4 allele (35.6% heterozygous and 7.2% homozygous); ApoE genotyping was not available for the healthy volunteers.

### 3.5. Risk prevalence across diagnoses

There was a statistically significant difference in age across diagnoses (F(3, 363) = 20.2, p<.001). Bonferroni post hoc analysis revealed that patients with a dementia diagnosis (mean=80.2, n=150) were significantly older than healthy volunteers (mean=76.4, n=81, p<.001), patients with an MCI diagnosis (mean=76.0, n=77, p<.001), and those who received non-memory disorder diagnoses or no formal diagnosis (mean=74.0, n=59, p<.001). There was also a statistically significant difference in the ACE-III total score across diagnostic groups (H(3)= 246.1, p<.001); post-hoc comparisons showed all interactions were significant.

There was a significant difference in depressive symptoms across diagnostic groups (H(3)= 16.38, p<.001), with lower total PHQ9 scores in healthy volunteers (mean = 2.9) than in those who received no dementia-related diagnosis (mean = 6.2). Physical activity also differed significantly across diagnoses (H(3)= 15.29, p=.002), driven by significantly more activity reported by healthy volunteers compared to patients with a dementia-related diagnosis. Patients with a diagnosis of MCI had a significantly greater BMI (mean = 26.9) than patients with a dementia-related diagnosis (mean = 24.9; H(3)= 10.09, p=.01), and healthy volunteers and patients with no memory-related/no formal diagnosis reported significantly less sleep than patients with a dementia diagnosis (H(3)= 20.91, p<.001).

Long-term conditions (LTCQ-8) total score also differed significantly (H(3)= 35.85, p<.001); healthy volunteers (mean = 85.7) scored significantly higher than patients with no memory-related/no formal diagnosis (mean = 77.9), MCI diagnosis (mean = 73.1), or a dementia-related diagnosis (mean = 69.6), and patients with no memory-related/no formal diagnosis scored significantly higher than patients with a dementia-related diagnosis. There was no significant difference in the prevalence of high-risk factors across diagnoses for education (H(3)= 5.15, p=.16) or alcohol consumption (H(3)= 7.43, p=.06), likely due to the small prevalence of these risk factors in all groups. There was no significant difference across diagnoses for the presence of an ε4 allele in patients (H(2)= 1.10, p=.58) or family history of dementia (H(3)= 1.61, p=.66). Figure 3 summarises the potentially modifiable risk for each diagnostic group: DRD, MCI, no DRD and healthy volunteers.

**Figure 3.**
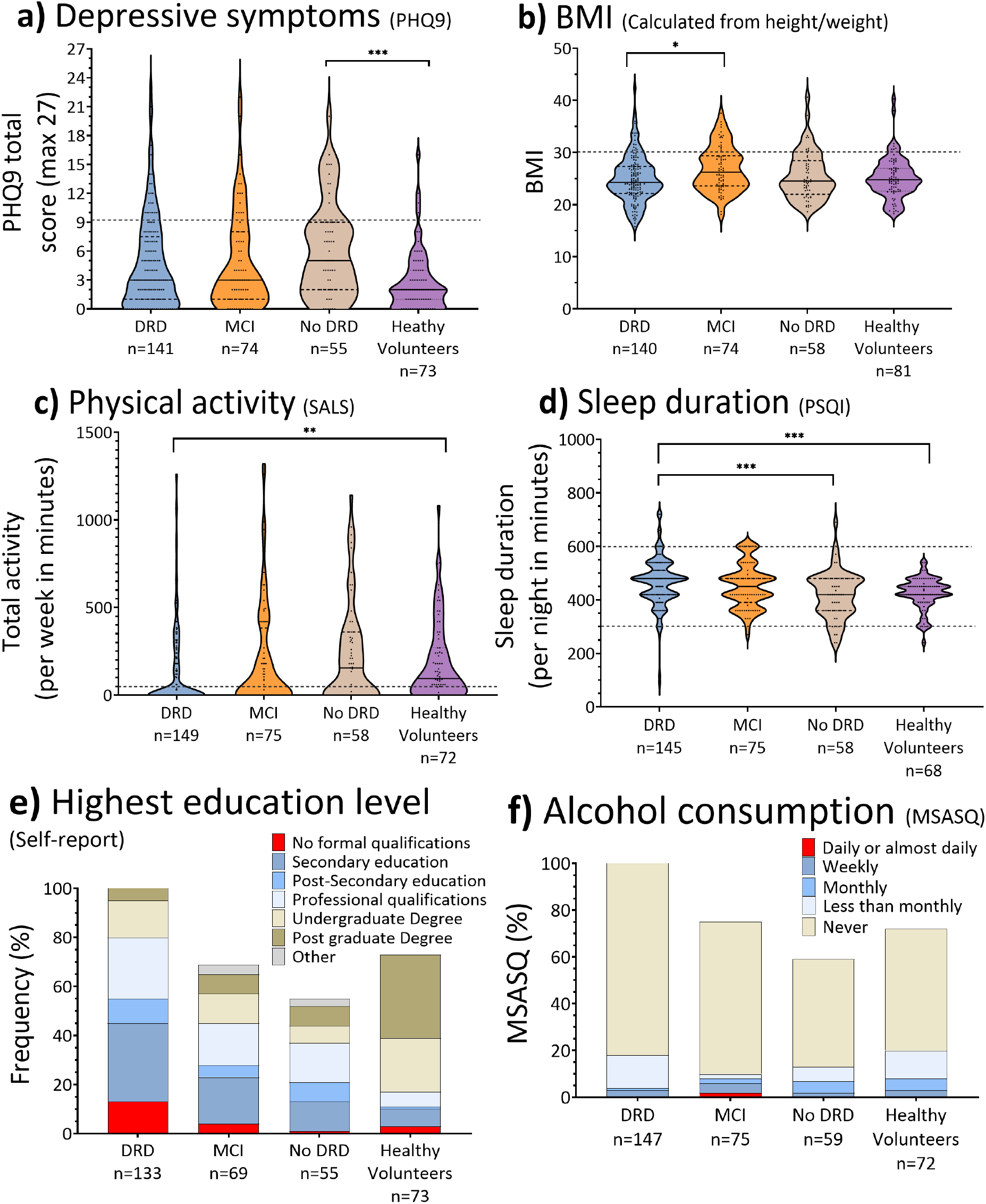
Prevalence of potenfially modifiable demenfia risks factors across OBHC diagnosis and healthy volunteers. The DRD (demenfia-related diagnosis) group consists of OBHC pafients who received a diagnosis with ICD-10 code: F00, F01, F02, F03; the MCI group consists of OBHC pafients who received a diagnosis with ICD-10 code F06.7, and the no DRD group consists of the OBHC pafients who received a non-memory disorder diagnosis e.g. primary psychiatric diagnosis (F31, F32, F41) or received no formal diagnosis after aftending the memory clinic. Dashed lines represent the high-risk thresholds.

### 3.6. Total number of potentially modifiable risks

67.4% of OBHC patients have one or more potentially modifiable dementia risk factors compared to 37.1% of healthy volunteers. This difference in total risk prevalence between groups was statistically significant (W = 17350, p<.001, effect size = -0.32), with OBHC patients having a greater number of risks (mean = 1.0 ± 0.6, range: 0-4) than healthy volunteers (mean = 0.4 ± 0.9, range: 0-3). There was also a significant difference in the total number of potentially modifiable dementia risk factors across diagnoses (H(3)= 32.64, p<.001), with healthy volunteers having significantly fewer risks (mean = 0.4) than patients with no dementia-related diagnosis (mean = 0.9), MCI (mean = 1.1) or dementia-related diagnosis (mean = 1.1). There was also a significant difference in cognition when the total number of potentially modifiable risks were grouped into 0, 1 and 2+ risks, with a significantly higher ACE-III total score in individuals with no risks (n= 147, mean = 84.5 ± 14.2, range: 23-100), than in those with one (n= 150, mean= 76.8 ± 18.4, range: 9-99) or 2 or more risks (n= 84, mean = 73.0 ± 19.3, range: 20-99). Figure 4 summarises the total number of potentially modifiable risk factors (education, physical activity, depression, BMI, sleep, alcohol consumption) for patients with a dementia-related diagnosis (a), MCI and no dementia-related diagnosis combined (b) and healthy volunteers (c) and the total ACE-III score across risk groups.

**Figure 4.**
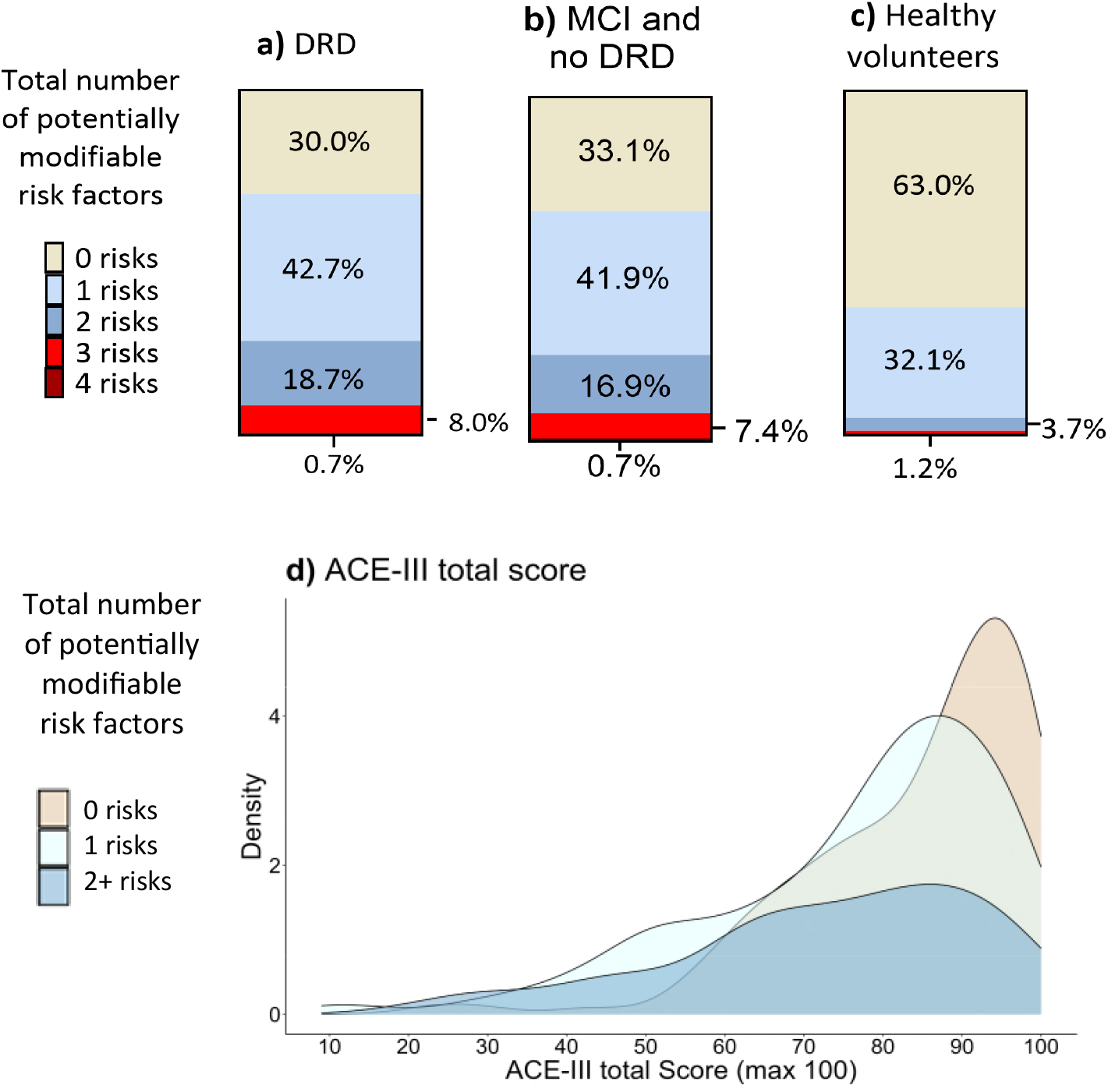
Total number of potenfially modifiable risk in OBHC pafients with a) a demenfia-related diagnosis, b) a diagnosis of MCI or no-DRD (non-memory disorder diagnosis or no formal diagnosis) and c) healthy volunteer; and (d) distribufion of cognifive scores across total number of potenfially modifiable risk factors. DRD = demenfia related diagnosis, ACE-III = Addenbrooke’s Cognifive Examinafion III.

## 4. Discussion

In this study, we described the demographic, cognitive, imaging, and diagnostic characteristics of patients in the OBHC population as well as the prevalence of potentially modifiable risk factors. We found that 91.5% (n=313) of patients who attended the OBHC consented to join the research database, resulting in a representative cohort of the population. Our findings demonstrate a significantly greater prevalence of potentially modifiable dementia risk factors in OBHC patients than in healthy volunteers. A total of 67.4% of OBHC patients have one or more potentially modifiable dementia risk factors and may be suitable for lifestyle interventions that target these risk factors. Furthermore, on the basis of the Lecanemab trial eligibility criteria, 24.6% of patients in the OBHC population may be eligible for further screening into DMTs.

This study provides an overview of the characteristics of the OBHC patient population in comparison to healthy volunteers and the average memory clinic population based on a 2023 audit^5^ in relation to age, demographics and diagnosis. The demographic characteristics of the OBHC population were similar to those of national memory clinic patients; the OBHC mean age (77.9 years) was two years lower than the average clinical population according to the MAS audit (79.7 years), and there was approximately 4% difference in the proportion of English as first language, where the first language is known. While the OBHC cohort had a lower proportion of patients with dementia-related diagnoses than the MAS audit cohort did, the most prevalent diagnoses of the OBHC cohort were dementia-related diagnoses, followed by MCI and non-memory disorder diagnoses and/or no formal diagnosis, in line with the MAS audit findings. The difference in prevalence rates is likely a result of the OBHC triage process, in which individuals with well-established and advanced cognitive impairments or individuals who are very frail are not referred to the OBHC. Although the triage process results in some selectivity, by integrating research into clinical practice, the OBHC has developed a highly inclusive cohort representative of the memory clinic patient population, driven by high research consent rates.

To the best of our knowledge, this study is one of the first to investigate the prevalence of potentially modifiable dementia risk factors in a real-world memory clinic population. We found a significantly greater prevalence of potentially modifiable dementia risk factors in OBHC patients than in healthy volunteers, and the results suggest that the majority of patients (67.4%) could benefit from lifestyle interventions to target these risk factors if evidence accumulates such that lifestyle interventions are developed. As expected, we observed a greater prevalence of all dementia risk factors in OBHC patients compared to healthy volunteers, which was significantly different for depressive symptoms, inappropriate sleep duration and inactivity. Patients with a dementia-related diagnosis also had a significantly greater prevalence of dementia risk factors compared to healthy volunteers and reported significantly less activity and sleep. These findings suggest that patients may benefit from interventions that particularly focus on improving mood, sleep, and physical activity. When we looked specifically at patients with a diagnosis of MCI or non-memory disorder/no formal diagnosis, we found that 66.9% of patients had at least one potentially modifiable risk factor, which was significantly greater than in healthy volunteers; these patients may benefit from lifestyle interventions targeting obesity and depression. As these patients may have fewer symptoms of cognitive impairment, lifestyle interventions that maximise the potential to delay or prevent dementia onset are crucial.

The findings also suggest that 24.6% of OBHC patients could potentially be eligible for further investigation into DMTs, specifically, Lecanemab, on the basis of trial eligibility criteria and findings from OBHC assessments, memory clinic diagnoses and radiology reports. The detailed imaging and non-imaging assessments completed at the OBHC allow for approximately three-quarters of patients to be ruled out from further screening, limiting unnecessary expensive and invasive investigations into amyloid status with PET or CSF assessment.

A few methodological considerations are relevant when interpreting this data. First, it is not possible to interpret the cause and effect of risk prevalence. The 2024 Lancet Commission on dementia^3^ updated the majority of potentially modifiable dementia risk factors, including alcohol consumption and obesity, to midlife risk factors, that affect later risk of dementia between the ages of 45-65 years. However, this study collected data on current risk prevalence only. Consequently, any potential lifestyle modifications may not have the same impact on patients at the time of admission to the OBHC as midlife interventions may. In addition, many risk factors, such as depressive symptoms and physical inactivity, may also present as symptoms of dementia, and without retrospective data, it is not possible to know when these risks began.

Second, while the OBHC population is highly representative of the local memory clinic population due to high consent rates, it has limited diversity compared with other areas of the UK. This is evidenced by the greater proportion of patients with white ethnicity than the national MAS rates. Dementia risk factors often disproportionately impact those with lower socioeconomic status, but we have no known information on the socioeconomic status of this cohort. These results also highlight the inherent difficulty in defining an adequate control group. As above, the healthy volunteer population may not be representative of the general population. The “healthy volunteer” bias suggests that people who volunteer for research are typically healthier than people who do not. For example, a greater proportion of healthy volunteers have advanced education (undergraduate and postgraduate) than may typically be expected in the general population. Development of a more diverse volunteer base is a well-known problem, and significant efforts are now being put into community engagement to reduce this bias in future research.

Finally, risk factor inclusion for this study was based on 6 of the 14 potentially modifiable risk factors identified in the Lancet Commission^3^ because risk data were not available for the clinical population for hearing loss, vision loss, traumatic brain injury, hypertension, high LDL cholesterol, smoking, social isolation, air pollution and diabetes. This may limit the interpretability of the total risk prevalence. Moreover, most risk data collected were based on self-report questionnaires, such as alcohol consumption, depressive symptoms, and physical inactivity, and OBHC patient family history of dementia was collected from medical records, where it was not consistently recorded and often unclear whether patients were reporting 1^st^- or 2^nd^-degree relatives. A number of ‘ideal’ and ‘practical’ procedures for risk data collection have been suggested by the European task force for BHS^9^; this study employed the ‘practical’ tool of self-reported alcohol consumption and physical activity as opposed to the suggested ideal tool of quantity-frequency measures with beverage-specific assessment of time frames and binge-drinking episodes, and accelerometers and smartwatches/phones (respectively). Consequently, these self-report methods would benefit from clinical validation. Data related to most of the missing risk factors are currently being extracted from patient medical records (for those who consent), and future studies with OBHC patients should consider the inclusion of missing risk factors and ‘ideal’ data collection tools, both for research and for developing good clinical advice.

In conclusion, this study comprehensively characterised the OBHC cohort and demonstrated that by integrating research into clinical practice, the OBHC has developed a cohort representative of the memory clinic patient population, driven by high research uptake. This study is one of the first to investigate the prevalence of potentially modifiable dementia risk factors and DMT eligibility in a real-world patient population. Approximately a quarter of patients would be suitable for further investigation into DMTs, and the majority of patients could potentially benefit from lifestyle interventions to address dementia risk factors. Understanding risk prevalence in clinical populations is essential for identifying high-risk groups and should inform the development of personalised risk reduction strategies to delay or prevent disease progression.

## 5. Data Availability

The data presented in this paper will be available via the Dementias Platform UK (https://portal.dementiasplatform.uk/CohortDirectory/Item?fingerPrintID=BHC) and access will be granted through an application process, reviewed by the BHC Data Access Group. The BHC Data Access Group will start accepting applications to access BHC data upon publication of the present work. Data will continue to be released in batches as the BHC progresses to minimise the risk of participant identification.

## 6. Acknowledgements

We are grateful to the operations team of the BHC (see https://www.psych.ox.ac.uk/research/translational-neuroimaging-group/team/oxford-brain-health-clinic).

## 7. Author contributions

**Jasmine Blane:** Conceptualisation, Data curation, Formal analysis, Investigation, Methodology, Visualisation, Writing – original draft. **Grace Gillis:** Investigation, Project administration, Writing – review & editing. **Ludovica Griffanti:** Conceptualisation, Methodology, Resources, Funding acquisition, Writing – review & editing. **Robert Mitchell:** Investigation, Writing – review & editing. **Pieter M Pretorious**: Investigation, Writing – review & editing. **Jane Fossey** Conceptualisation, funding acquisition, Writing – review & editing. **Vanessa Raymont:** Conceptualisation, Resources, Funding acquisition, Supervision, Writing – review & editing. **Lola Martos:** Conceptualisation, Resources, Funding acquisition, Writing – review & editing. **Clare E Mackay:** Conceptualisation, Methodology, Resources, Funding acquisition, Supervision, Writing – review & editing.

## References

1 Alzheimer’s Research UK. Tipping point: the future of dementia. (2023).

2 Pittock, R. R. et al. Eligibility for Anti-Amyloid Treatment in a Population-Based Study of Cognitive Aging. Neurology 101, e1837–e1849 (2023). 10.1212/WNL.0000000000207770

3 Livingston, G. et al. Dementia prevention, intervention, and care: 2024 report of the Lancet standing Commission. The Lancet 404, 572–628 (2024). 10.1016/S0140-6736(24)01296-0

4 Long, S., Benoist, C., Weidner, W. World Alzheimer Report 2023: Reducing dementia risk: never too early, never too late. (London, England, 2023).

5 Royal College of Psychiatrists. National Audit of Dementia Spotlight Audit in Memory Assessment Services national report. (London: Healthcare Quality Improvement Partnership, 2023/24).

6 van der Flier, W. M. & Scheltens, P. Amsterdam Dementia Cohort: Performing Research to Optimize Care. J Alzheimers Dis 62, 1091–1111 (2018). 10.3233/jad-170850

7 Kivipelto, M. et al. The Finnish Geriatric Intervention Study to Prevent Cognitive Impairment and Disability (FINGER): Study design and progress. Alzheimer’s & Dementia 9, 657–665 (2013). 10.1016/j.jalz.2012.09.012

8 Ngandu, T. et al. A 2 year multidomain intervention of diet, exercise, cognitive training, and vascular risk monitoring versus control to prevent cognitive decline in at-risk elderly people (FINGER): a randomised controlled trial. The Lancet 385, 2255–2263 (2015). 10.1016/S0140-6736(15)60461-5

9 Frisoni, G. B. et al. Dementia prevention in memory clinics: recommendations from the European task force for brain health services. The Lancet Regional Health – Europe 26 (2023). 10.1016/j.lanepe.2022.100576

10 O’Donoghue, M. C. et al. Oxford brain health clinic: protocol and research database. BMJ Open 13, e067808 (2023). 10.1136/bmjopen-2022-067808

11 Cummings, J. et al. Lecanemab: Appropriate Use Recommendations. J Prev Alzheimers Dis 10, 362–377 (2023). 10.14283/jpad.2023.30

12 Hsieh, S., Schubert, S., Hoon, C., Mioshi, E. & Hodges, J. R. Validation of the Addenbrooke’s Cognitive Examination III in frontotemporal dementia and Alzheimer’s disease. Dement Geriatr Cogn Disord 36, 242–250 (2013). 10.1159/000351671

13 Smith, P. C., Schmidt, S. M., Allensworth-Davies, D. & Saitz, R. Primary care validation of a single-question alcohol screening test. J Gen Intern Med 24, 783–788 (2009). 10.1007/s11606-009-0928-6

14 Kroenke, K., Spitzer, R. L. & Williams, J. B. The PHQ-9: validity of a brief depression severity measure. J Gen Intern Med 16, 606–613 (2001). 10.1046/j.1525-1497.2001.016009606.x

15 Milton, K., Engeli, A., Townsend, T., Coombes, E., Jones, A. The selection of a project level measure of physical activity. (London: Sport England, 2017).

16 Batchelder, L. et al. Rasch analysis of the long-term conditions questionnaire (LTCQ) and development of a short-form (LTCQ-8). Health Qual Life Outcomes 18, 375 (2020). 10.1186/s12955-020-01626-3

17 Devlin, N., Pickard, S. & Busschbach, J. in Value Sets for EQ-5D-5L: A Compendium, Comparative Review & User Guide 1–12 (Springer International Publishing, 2022).

18 Bucks, R. S., Ashworth, D. L., Wilcock, G. K. & Siegfried, K. Assessment of activities of daily living in dementia: development of the Bristol Activities of Daily Living Scale. Age Ageing 25, 113–120 (1996). 10.1093/ageing/25.2.113

19 Griffanti, L. et al. Adapting UK Biobank imaging for use in a routine memory clinic setting: The Oxford Brain Health Clinic. NeuroImage: Clinical 36, 103273 (2022). 10.1016/j.nicl.2022.103273

20 Livingston, G. et al. Dementia prevention, intervention, and care: 2020 report of the Lancet Commission. The Lancet 396, 413–446 (2020). 10.1016/s0140-6736(20)30367-6

21 Matías-Guiu, J. A. et al. Conversion between Addenbrooke’s Cognitive Examination III and Mini-Mental State Examination. Int Psychogeriatr 30, 1227–1233 (2018). 10.1017/s104161021700268x

